# Understanding Disordered Eating Attitudes and Patterns in University Students and the Relationship to Campus Dining Services

**DOI:** 10.64898/2026.05.11.26352946

**Authors:** Benjamin Bartling

## Abstract

University Students are particularly vulnerable to disordered eating behaviors (DEB) and attitudes (DEA). This study expands upon the knowledge base of DEA and DEB in university students by employing a netnography as a precursor to the main study to establish the following research questions: What is the relationship between the perceived quality of dining services and DEA? What is the relationship between the perceived availability of dining services and DEA? And lastly, how does prior experience with dining services affect eating patterns and attitudes toward food? The first study utilized a netnographic approach in order to evaluate issues with university dining services, leading to the design of the second study. Students at an upper Midwestern university (*n*=88) were surveyed via convenience sampling. Eating attitudes, eating behaviors, and relationships with dining services were measured. A statistically significant relationship between the availability of services and DEA was found. A statistically significant relationship between the availability of services and risk behaviors was found. However, no statistically significant correlation existed between first-year dependence on on-campus dining services and risk behavior related to eating disorders or eating attitudes. Based on this, we know the quality of nutrition and the availability of services impacted students’ eating attitudes and behaviors, not inherent dependence.

## 1. Introduction

College is a critical period in the development of a student with respect to emotions, physical health, and academic success (Dvořáková et al., 2019). Students enter university with limited foresight into what their day-to-day lives will look like. Most importantly in this context, students must now find new avenues to fulfill their nutritional needs. Despite many universities mandating that students purchase a meal plan while living on campus, students also spend money on groceries and eat off campus, further increasing the financial burden of nutrition. As such, college students are susceptible to a range of adverse health outcomes due to insufficient knowledge of how to care for their bodies through nutrition (Sheldon et al., 2021).

Often, when young adults enter college, it is the first time they leave home, which allows them the freedom and responsibility to make their own decisions regarding their health and nutrition. This drastic change in environment and responsibility can lead to irregular eating patterns, thereby eliciting disordered eating behaviors and attitudes (Treasure et al., 2020). Some individuals are predisposed to disordered eating through their genetics, combined with cultural and environmental factors. Inherent vulnerability leaves certain population groups susceptible to disordered eating, primarily young adult college students. This concept of changes to the eating environment and disordered eating behaviors will be discussed later in this paper.

The impact of campus dining services on disordered eating behaviors and attitudes is a severely overlooked area of research. Additionally, there is still a lack of information regarding the specific mechanisms that lead to adverse outcomes. While it is evident that the quality and availability of dining services play a role, it remains unclear how these factors interact with individual differences in genetics, culture, and personal experience to produce problematic eating behaviors and attitudes. Therefore, there is an urgent need for further research that delves deeper into the causal mechanisms that underlie the relationship between campus dining services and disordered eating behaviors and attitudes among university students.

## 2. Literature Review

### 2.1 Conceptualizing Eating Disorder Etiology

There is a great deal of ambiguity surrounding the conceptualization of eating disorder etiology and disordered eating behaviors. Should eating disorders be attributed to eating patterns and body image, or are they psychotic, psychosomatic, or neurotic disorders (O’Brien and Vincent, 2003)? The clinical presentation of an eating disorder as it progresses with age becomes increasingly complex (Nakazato et al., 2012). Thus, eating disorder pathology will likely differ from childhood and adolescence to later stages in the adult developmental cycle. Understanding the interplay between eating disorders and developmental processes is crucial to identifying disorders at different life stages, as well as the treatment and maintenance of these disorders.

In addition, the interaction between genetics and environmental stressors during critical developmental periods adds to the complexity of properly modeling these disorders. Environmental factors are difficult to account for, but it is crucial to consider these factors in order to have a comprehensive understanding of the etiology of eating disorders and disordered eating behavior. Pike et al. (2021) found risk factors for AN and BN in Japan. These include body image disturbance, peer and family factors, and media exposure. They argue that the steady accumulation of stressful events in the year immediately preceding the onset of disturbances to eating behaviors indicated increased risk.

Lengthy periods of untreated eating disorder symptoms can lead to an ingrained form of the disorder within the person, which makes it increasingly more difficult to treat (Potterton et al., 2021). Eating disorders are serious psychiatric illnesses and are characterized by abnormal eating or weight control behaviors. Distorted attitudes towards eating, weight, and body shape play a critical role in the origin and development of eating disorders.

There are six main eating and feeding disorders that are recognized by the American Psychological Association, however, AN, BN, and BED are the main three. The Diagnostic and Statistical Manual of Mental Disorders (5th ed.; DSM-5; American Psychiatric Association, 2013). is the most universally accepted wording used by researchers and clinicians all around the world for the classification of mental disorders. The DSM-5 also provides subtype qualifiers for certain disorders like BN and AN. These subtype disorders do not meet the threshold for a clinical diagnosis however they are indicators for the severity of the disorder, and help lead clinicians to clear, concise definitions of remission.

In the population of individuals suffering from an eating disorder, psychiatric comorbidities are exceptionally common, with rates surpassing 70% for any two or more mental health disorders. Additionally, the most frequently seen comorbidities include substance abuse, personality disorders, and anxiety disorders (Treasure et al., 2020). Having an eating disorder leaves a person susceptible to a plethora of issues and problems, both mentally and physically. Proper treatment and diagnosis of eating disorders and disordered eating patterns are crucial to alleviating the suffering these psychiatric conditions inflict on the individual and those in their immediate circle of family and friends.

Additionally, the concept of food insecurity plays a significant role in the development and perpetuation of disordered eating behaviors and attitudes and must be understood in order to grasp the full magnitude of this study.

### 2.2 Food Insecurity

Food insecurity is the limited, uncertain, or unreliable access to a sufficient quantity of nutritionally adequate food needed to sustain proper health (Christensen et al., 2021). University students are especially vulnerable to the effects of food insecurity due to the massive demands placed upon their financial resources (Archibald and Feldman, 2018).

Food insecurity in university students is especially detrimental because it is linked to poorer health outcomes including subpar physical activity, low nutrition, quality diet, obesity, anxiety, and depression. Unfortunately, this means that 39% of university students, roughly 8 million young adults, in the United States experienced food insecurity at some point during the fall semester of 2019 (Royer et al., 2021).

Alternating periods of access to food and deprivation mimics the physiological and behavioral effects of dieting and may promote overeating behaviors mainly due to elevated periods of consistent hunger deprivation (Becker et al., 2019). Dieting behaviors increase the likelihood of an individual developing a binge eating disorder due to extended periods of physiological hunger. In this case, binge eating further reinforces the perception of the need to diet in order to compensate for calories consumed during an episode. This is especially evident in cultures, mainly western, that emphasize thinness (Becker et al., 2019). Individuals who suffer from food insecurity are at a significantly higher risk for developing an eating disorder. Indeed, research has found that 17% of individuals suffering from food insecurity meet the clinical threshold for a diagnosis of an eating disorder (Becker et al., 2017). Additionally, a diagnosis of bulimia nervosa and binge eating disorder were much more common in food insecure populations vs food secure populations (Lydecker and Grilo, 2019).

Additionally, food-insecure first-year college students reported significantly higher disordered eating behaviors than food-secure students (Goldrick-Rab, 2016). This illustrates a very concerning trend showing food insecurity and disordered eating behaviors worsening concordantly. This trend suggests that students experiencing food insecurity likely have coexisting disordered eating behaviors that may only be ceased if food security improves, alluding to a systemic issue (Royer et al., 2021). The financial burden placed upon students by universities must be met to continue with the pursuit of higher education. These high-priority demands must take precedence over other needs, such as consuming nutritionally adequate food (Goldrick-Rab et al., 2017). This leads to lower food budgets, reduced food intake, and consumption of inexpensive energy-dense foods like soft drinks and fast food that also have low nutritional value. These energy dense foods are considerably like the nutritional services offered by universities for students who live on campus (Bruening et al., 2012).

Increasing barriers to food access, such as lower food availability, lower quality of food available for purchase, loss of access to food programs, and increased food for delivery, may exacerbate the issue of food insecurity in university students.Christensen et al. (2021) found that 47.6% of individuals with food insecurity scored positive for an ED. This study is important because it demonstrates the relationship between food insecurity and an ED diagnosis, thus alluding to a high number of disordered eating behaviors specifically in the population of university students.

### 2.3. College Dining Services

University dining halls offer a wide variety of food choices. However, there is minimal information available for university students at the point of selection in all-you-can-eat campus dining halls, where the cost of food is controlled (Freedman, 2011). Pilot projects conducted with vending machines (Wilbur et al., 1978) and in school cafeterias (Zifferblatt et al., 1978) found that presenting nutritional information at the point of purchase had the potential for producing significant changes in nutrition consumption. This effect was strongly replicated in supermarket and similar settings (Pohl and Freimuth, 1983).

The average meal plan costs roughly $4,500 for a twosemester academic year (DeAmelio-Rafferty, 2022). This means a student would have roughly $18.75 a day to spend on food with a university meal plan. That is 70% more per day to eat on campus, compared to the $11 a day the Bureau of Labor Statistics (2022) estimates a single person spends on food. What this means is that college students are essentially paying more to eat than the average American. According to Sheldon et al. (2021), it is implied that it is the university’s responsibility to provide adequate nutrition for students. There are limited options on university campuses, and this leaves students with minimal choices and subpar, heavily processed food.

The reality of being forced to learn how to make the appropriate nutritional choices once entering university is quite difficult for many students. Easily accessible items such as fast food and store-bought preheatable meals are becoming increasingly common among college students, replacing home-cooked meals. This is increasingly more prevalent in students taking a full semester’s worth of credits. Dietary habits for many college students do not meet recommended guidelines set out by healthcare professionals, causing weight to increase in both males and females from their first year of college to when they graduate (Mokdad et al., 1999). The obesity rate among university students in the United States increased from 12% to 36% of the population from 1991 to 2004 (Ogden and Carroll, 2006). The American College Health Association showed a similar trend in 2010 at a rate of 33% (American College Health Association, 2011). This data shows how crucial nutrition is for young adults and its importance in higher education.

Establishing healthy eating and exercise patterns is crucial to prevent disordered eating behaviors.Huber et al. (2021) found that a massive interruption caused by the COVID-19 pandemic to the food environment had an immediate and significant impact on food procurement and dietary habits in young adults. The data showed that government lockdowns drastically altered the amount of food individuals consumed. Specifically, 16.8% of respondents indicated reduced daily intake, while 31.2% indicated an increase in food consumption. This data is important because it shows that a major change to the food environment drastically affected the eating behaviors of 48% of participants. The amount of food consumed was highly influenced by the individual’s gender, body mass index, physical activity, smoking habits, psychological stress, and alcohol consumption (Huber et al., 2021). Proper nutrition is the first line of defense against a number of complex disordered eating behaviors and attitudes.

### 2.4. Eating Attitudes

A conceptual definition for eating attitudes is the thoughts, feelings, beliefs, behaviors, and relationships regarding food an individual possesses (Alvarenga et al., 2008). Being able to understand these attitudes is crucial to help develop reliable and consistent interventions based on food choices and nutritional counseling. Changing disordered eating attitudes is essential for the proper treatment of eating disorders.

There are two types of eating attitudes an individual possesses: explicit and implicit. Explicit eating attitudes can be defined as consciously accessible mental processes. Implicit attitudes encompass past experiences with food, accompanied by the inability to reflect consciously, that have an impact on cognitive patterns, actions, and behaviors toward a mark (Nosek et al., 2012). Explicit and implicit eating attitudes are distinct categorical constructs from one another. However, they may be the same or similar in individuals.

Explicit attitudes can be attributed to a number of different factors, such as intention, social desirability, aptitude, and normative attitudes (Czyzewska and Graham, 2008). It is important to note that explicit eating attitudes may not be the most predictive of eating behavior. However, research suggests that implicit attitudes and other forms of automatic processes strongly influence food selection choices (Cervellon et al., 2007). Previous literature has demonstrated the attitude-behavior relationship as highly complex and reciprocal, which essentially means that attitudes influence behavior in a bidirectional relationship (Dorsey et al., 2009).

Compared to healthy individuals who do not present with any ED symptoms, people with ED symptoms are exceedingly more likely to hold the belief that having an eating disorder is acceptable or desirable (Mond and Arrighi, 2011). Individuals with AN highlight the disorder’s perceived benefit, including avoidance of negative emotions, a heightened sense of internal locus of control, and the irrational fear of losing these perceived benefits as a barrier to seeking treatment (Bullivant et al., 2020). For AN and BN, several general eating attitudes have been described including incompetence in dealing with meals, eating with others, difficulty with making food choices (American Dietetic Association, 2006), bifurcated classification of food as “good” or “bad,” experiencing anger in response to hunger sensations and cues (Sunday et al., 1992), and using food to cope with negative emotions and distorted beliefs about the nutritional value of food (American Psychiatric Association, 1994). For binge eating disorder patients, eating attitudes encompass feelings of high loss of control, chaotic eating habits, and similar maladaptive behaviors (Ellis et al., 2020).

Recent research suggests that the increasing incidence of eating disorders can be attributed to changes in the food environment (Treasure et al., 2020). The food environment can be defined as the economic, physical, and socio-cultural contexts and situations in which people engage with food. This research is crucial when contextualizing the relationship between campus dining services and disordered eating attitudes and behaviors in university students.

This study sought to better understand the relationship between the perceived quality and availability of dining services and disordered eating behaviors and attitudes in university students. To achieve this, the primary author employed an inductive research strategy in the form of a netnography as a precursor to the main study in order to warrant further investigation. The netnographic approach was used to evaluate issues with university dining services primarily via the outspoken concerns from community members and subsequent interactions, leading to the design of the second study.

The second study involved surveying students at an upper Midwestern university via convenience sampling as well as other sampling methods such as snowball sampling, posters, and administrative campus email lists. Eating attitudes, eating behaviors, and relationships with dining services were measured to confirm the findings of the inductive research strategy. Given all this information, we will investigate this relationship by analyzing the sentiment of students’ eating attitudes and behaviors toward campus dining services through the following research questions:

RQ1: How does satisfaction with campus dining services affect university students’ eating patterns and attitudes toward food?

RQ2: What is the relationship between the perceived availability of campus dining services and disordered eating attitudes in university students?

RQ3: What is the relationship between perceived quality of campus dining services and disordered eating attitudes in university students?

## 3. Study 1: Longitudinal Netnography

This first exploratory study utilized an online ethnographic approach to gain insight into students’ perceptions of campus dining services and general eating attitudes or behaviors. Netnography is a qualitative research method that is analogous to ethnographies that immerses researchers in digital communities in order to contextualize available online data (Kozinets, 2010). This research method offers researchers rich qualitative data and is a useful tool for collecting and analyzing information from a wide variety of people because of its accessibility.

Netnography is based on an ethnographic research approach to study and understand consumers’ thoughts and attitudes toward a specific topic (Kozinets, 2006). Netnography is a distinct research method that utilizes its own set of methodological guidelines (Caliandro, 2014).Kozinets (2010) recommends four procedures or methodological stages for netnographic studies. These stages include entree, which is the formation of a research hypothesis and distinguishing a web-based population to research. Next, data collection is the direct communication from community members on a web platform regarding its interactions and meanings. Lastly, analysis and interpretation involve conceptualizing, coding, and classifying communicative interactions (Bowler, 2010).

With the increasing advancement of technology, individuals constantly interact with online platforms before, after, and during their use of services, creating a unique set of information regarding their activities and experiences. This unique set of information is naturally occurring and unprompted, thereby providing relatively unbiased qualitative data. A more involved and active approach to netnographic studies has been found to significantly facilitate offline research methods such as observations in two major ways. First, netnographies increase social and cognitive proximity between researchers and participants. Second, they enable the expanded spatial and temporal network of a researcher’s fieldwork (Wu, 2022).

A netnographic approach was selected for use in this research project because of its accessibility, anonymity, and convenience to gauge student attitudes and perceptions toward campus dining services, as well as the overall eating culture at an upper Midwestern university through its use as an exploratory research tool. This research method allowed the researcher to unobtrusively view the natural communication and cognitive processes about dining services that were believed to be directly related to disordered eating behaviors and attitudes in university students.

The online web platform known as Yik Yak is an anonymous application that allows its users to connect with others within five miles as well as students who attend the same university. This makes it particularly enticing for college students looking for a sense of community and a space to vent their frustrations and opinions. On the application, every user must authenticate their identity with a valid United States phone number. Every user’s identity is kept confidential, allowing users to express their opinions and share information freely. Users are able to freely post, comment on other posts, and upvote or downvote content. Upvoting is the application’s method of liking or agreeing with a post.

The utilization of Yik Yak in this netnography allowed the researcher to perform an exploratory investigation into the perceptions of university students at an upper Midwestern university regarding campus dining services and the quality and availability of nutritional services.

### 3.1. Methods Study One

This netnography employed a grounded theory approach in the collection of qualitative data (Glaser, 1978; Strauss and Corbin, 1990). A grounded theory approach was selected because it allows for an understanding of overarching themes that contribute to a theoretical framework. Yik Yak was utilized from August 2021 to December 2022.

Over this time period, twenty-two posts were documented illustrating students’ frustrations and negative attitudes toward on-campus dining services at an upper Midwestern university for the scope of this research, although considerably more posts manifested similar themes. In a typical week there are upwards of 1,250 unique posts with over 700 comments. Topics may include dorm life, academics, social events, or, in the interest of this study, campus dining services.

As an open forum platform, Yik Yak allows for the instantaneous and spontaneous recounting of the lived experiences of university students. When comparing the number of posts related to campus dining services and eating behaviors to the total number of posts in a certain time frame, it is critical to demonstrate that while there may not be a vast number of posts regarding eating behaviors and sentiment toward campus dining services, these topics nevertheless appear important to students because of the shortand long-term effects on their mental and physical health.

In this study, a grounded theory approach allowed the researcher to analyze the lived experiences of students’ relationships with campus dining services from multiple perspectives.

### 3.2. Results Study One

#### 3.2.1. Theme 1: Negative Perceptions Toward Nutritional Services

There appears to be a continued and important relationship between student perceptions of dining services and eating attitudes or behaviors. This theme illustrates the relationship between campus dining services and university students’ disordered eating behaviors and attitudes.

One individual posted:

“Can my ED [eating disorder] go away this is just annoying now.”

As netnographies allow for the researcher to be an active participant (Kozinets, 2010), one post asked:

“Comment/Upvote if you believe that campus dining led you to develop an eating disorder or worsened your already existing ED.”

The responses were overwhelming, with over fifty upvotes after 14 hours, whereas a typical post receives approximately three to four upvotes. Beyond upvotes, responses to the post included:

“So it wasn’t just me [referring to disordered eating because of campus dining].”

Additional comments included descriptions of altered eating habits such as:

“I eat a spoonful of Nutella a day.” “I never eat… It’s so bad.”

The numerous posts that conveyed a similar message led to the creation of this theme regarding the relationship between campus dining services and disordered eating attitudes or behaviors. The volume of responses to this post was especially concerning when considering the context of students being under considerable pressure to succeed academically and socially, which may contribute to disastrous consequences in the long run.

#### 3.2.2. Theme 2: Health Concerns Regarding Perceived Nutritional Quality

This theme comprises students’ health concerns regarding the perceived quality of nutrition they are receiving from campus dining services. After witnessing overwhelmingly negative feedback about on-campus dining services, the primary researcher began collecting various posts from Yik Yak that demonstrated students’ sentiment regarding their health concerns with campus dining services.

Examples included:

“Is the [student dining service] food giving anyone else diarrhea?”

Comments included:

“Absolutely.”

“Hershey squirts are tough right now.” Another post stated:

“Anyone else have absolutely terrible shits from [student dining services] food?”

This post received eight upvotes within an hour, with comments including:

“Straight from the bowels of hell…” “Anything I eat in there, swear to god.”

Additional comments included:

“The [student dining services] food gives me the shits 0/10.”

“[Student dining services are] OBLITERATING my stomach.”

“It makes me sick every time I eat it, but it’s that or starve.”

“[Student dining services] food this week has got my stomach acting up.”

“This week, my stomach hates the [student dining services] food.”

One fast-food Mexican restaurant appeared to be particularly concerning for students’ health and possessed a notably poor reputation on campus, as illustrated by the following statements:

“Swear to god [Mexican fast-food restaurant] gave me food poisoning.”

“PSA: got food poisoning from [Mexican fast-food restaurant] and been yaking shit up ever since I ate there. Looking out for y’all.”

“Anyone else get sick EVERY time they eat [Mexican fast-food restaurant] from the [student dining center]?”

“Hair in my [Mexican fast-food restaurant].”

The general theme these posts conveyed over the course of the year was that students were extremely frustrated and upset with the quality of food provided by campus dining services and the effects it had on their digestive systems and overall health.

There are some limitations related to netnography and the data it produces. A primary concern is the authenticity and quality of the data collected (Xun and Reynolds, 2010). There is also difficulty establishing demographic information from the collected data. However, for the purposes of this study, quality of the data collected was moderated by the careful selection of student responses while maintaining neutrality in order to avoid eliciting false responses.

### 3.3. Discussion Study One

Based on the netnography, significant themes were identified. Theme 1 comprised the appearance of a continued and important relationship between student perceptions of dining services and eating attitudes or behaviors. This led to the conclusion that campus dining services may play a role in the development, maintenance, and progression of disordered eating patterns and attitudes in university students.

This conclusion raised the following questions:

How does prior experience with campus dining services affect university students’ eating patterns and attitudes toward food?

Is there a statistically significant relationship between the perceived availability of campus dining services and disordered eating attitudes in university students?

Theme 2 comprised students’ health concerns regarding the perceived quality of nutrition they receive from campus dining services. This theme led to the conclusion that students are discontent with the quality of nutrition available from on-campus dining services and the subsequent digestive issues.

This conclusion raised the following question:

Is there a positive relationship between students’ eating patterns and attitudes toward food in general and perceived quality of on-campus dining services?

These questions became the focus of the second part of this mixed-methods research study. The second study was necessary because very little research has investigated university dining services and how those services affect students’ eating attitudes and behaviors in both the short and long term. Thus, the relationship between the perceptions of university students and campus dining services is a severely under-researched area of literature requiring further investigation.

This is crucial in order to further comprehend the issue of disordered eating behaviors and attitudes in university students and to provide the foundation for the second research project. The thematic coding of qualitative data from the netnography allowed the researcher to identify a significant relationship between perceptions of campus dining services and students’ eating attitudes and behaviors.

This study aims to identify a statistically significant relationship between campus dining services and students’ eating attitudes and behaviors. The findings provide future research with a foundation to further investigate the issue of perceived availability and quality of nutritional services provided by universities and the relationship to disordered eating behaviors and attitudes in university students.

Based on the previous study and the noticeable gap in the current literature, the following hypotheses were proposed:

H1: There is a positive relationship between perceived quality of on-campus dining services and disordered eating attitudes in university students.

H2: There is a positive relationship between perceived availability of on-campus dining services and disordered eating attitudes in university students.

## 4. Study 2

### 4.1. Methods Study Two

#### 4.1.1. Procedures

Following informed consent, participants were asked to complete the survey by answering questions regarding the variables explained below. Surveys were completed online via Qualtrics. Once participants completed the survey, they read a debriefing statement and were provided with mental health crisis resources.

#### 4.1.2. Participants

A total of 121 undergraduate students at an upper Midwestern university participated in this study and were offered the chance to win one of twenty $35 electronic gift cards. Participants were recruited through social media, flyers, and the university’s honors email list.

Participants ranged in age from 18 to 24 years old (*M* = 19.91, *S D* = 2.11) and were primarily female (85%). Additionally, participants were primarily Caucasian (84.1%). This population was selected because university students are highly susceptible to disordered eating attitudes and behaviors through a combination of environmental, social, and biological factors (Treasure et al., 2020).

Any participant who completed less than 70% of the survey was removed from the study. This resulted in a final sample of *N* = 88 individuals.

#### 4.1.3. Measures

*Disordered Eating Attitudes Scale (DEAS)*. The DEAS questionnaire (Alvarenga et al., 2010) evaluates beliefs, feelings, thoughts, behaviors, and relationships with food. This scale has been used for screenings of nutritional deviations in nonclinical populations (Alvarenga et al., 2010). The DEAS questionnaire consists of 25 items measured on a 7-point Likert-type scale.

Illustrative examples include:

“I dream of a pill that would replace food.”

“I worry about how much a certain kind of food or meal will make me gain weight.”

The original scale demonstrated the following psychometric properties (*M* = 1.87, *S D* = .34, α = .22).

Components of the DEAS scale were used to create the DEARC measure, which evaluates an individual’s attitudes and behaviors regarding their relationship with food, concerns about food and weight, and subjective feelings toward eating.

The revised measure demonstrated acceptable internal reliability (α = .77).

The subscales utilized were subscales 1, 2, and 4. These subscales measured relationship with food, concerns about food and weight gain, and feelings toward eating. Subscales 3 and 5 measured restrictive and compensatory practices and ideas about normal eating; however, they reduced the reliability of the overall measure and were therefore removed despite individually demonstrating acceptable internal reliability.

DEAS subscale one (DEAS1), relationship with food, demonstrated acceptable internal reliability (α = .82) with three items.

Subscale two (DEAS2), concerns about food and weight gain, demonstrated acceptable internal reliability (α = .80) after item DEAS2.01 was removed, increasing reliability from α = .27 to α = .80 with three items.

Subscale three (DEAS3), restrictive and compensatory practices, demonstrated acceptable internal reliability (α = .78) after item DEAS3.04 was removed, increasing reliability from α = −.40 to α = .78 with three items.

Subscale four (DEAS4), feelings toward eating, demonstrated excellent internal reliability (α = .91) with three items.

Subscale five (DEAS5), ideas of normal eating, demonstrated acceptable internal reliability (α = .73) with fourteen items.

The complete Disordered Eating Attitudes Scale (DEAS) initially demonstrated poor overall reliability (α = .22). Removing DEAS3 and DEAS5 increased the overall reliability of the revised measure to α = .77.

*Risk Behavior Related to Eating Disorders (RiBED-8)*. Risk Behavior Related to Eating Disorders (RiBED-8) is an eight-item instrument designed to assess participants’ eating-related behaviors and attitudes on a scale from 1 to 4, with responses ranging from “Never,” “Seldom,” “Often,” and “Very Often.”

Examples of this measure include:

“I diet.”

“I have a bad conscience because I eat sweets.” “I throw up to get rid of the food I have eaten.”

The measure demonstrated acceptable psychometric properties (*M* = 1.92, *S D* = .52, α = .72).

The measure was specifically designed to capture risk behaviors associated with eating disorders and has been found effective in predicting clinical eating disorders. The scale demonstrated acceptable psychometric properties for girls (Waaddegaard et al., 2003).

*Satisfaction and Relationship with Campus Dining*. Students were asked questions regarding their dependence on on-campus dining services, such as receiving the majority of their meals from on-campus food providers, along with their thoughts, beliefs, and attitudes toward on-campus dining services using a seven-point Likert scale.

Questions regarding availability of services, nutritional quality, scheduling conflicts, energy levels, and illness were included to assess students’ relationships with campus dining services.

This scale was categorized as RCS and broken into the following dimensions:

**RCS1**. Dependence on on-campus dining services during the first year at university (*M* = 5.95, *S D* = 1.61, α = 1.00).

Perceived quality of nutritional services consisted of RCS4 and RCS6 (*M* = 4.66, *S D* = 1.56, α = .87).

Perceived availability of services consisted of RCS3, RCS5, and RCS7 (*M* = 4.88, *S D* = 1.35, α = .74).

RCS4 and RCS6 were combined to form the subjective quality measure (QUA; α = .87).

RCS3, RCS5, and RCS7 were combined to form the subjective availability measure (AVA; α = .74).

The overall measure was termed Satisfaction (SAT; *M* =

4.69, *S D* = 1.26, α = .80).

### 4.2. Results Study Two

Missing data points within variables were replaced with mean scores from the corresponding variable (*N* = 88). Missing values were replaced using mean substitution rather than invalid responses because there were no major outliers within the dataset, thereby preserving the mean while minimally reducing standard deviations (Lang & Little, 2018). Due to the relatively low number of missing data points, the impact on the standard deviation was considered negligible.

All items demonstrated acceptable internal reliability.

Given the multiple statistical comparisons utilized, a Bonferroni correction was employed to reduce the likelihood of Type I errors. Each test was designed using a significance threshold of *p* = .05 and a one-tailed *t*-test. Following the correction, the adjusted significance threshold became *p*_new_ = .016.

For the power analysis, a standard deviation of 1.13 was identified. A power level of 90% with a significance level of .05 was selected for this study. The standard deviation was derived from a set of community norms within the existing literature (*N* = 243; (Fairburn and Beglin, 1994). A minimum sample size of 54 participants was identified. The sample utilized in this study exceeded the required sample size, supporting sufficient statistical power.

The first hypothesis predicted that there would be a positive relationship between perceived quality of on-campus dining services and disordered eating attitudes in university students.

Directional *t*-tests with Bonferroni adjustments indicated that higher dissatisfaction (lower perceived quality) with nutritional services was associated with increased disordered eating attitudes and patterns.

A Pearson correlation coefficient was computed to assess the linear relationship between satisfaction (perceived quality) with nutritional services and disordered eating attitudes. There was a positive correlation between the two variables:

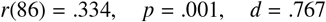

Additionally, a Pearson correlation coefficient was computed to assess the linear relationship between satisfaction with nutritional services and disordered eating behaviors. There was also a positive correlation between these variables:

There was a positive correlation between the variables (*r*(86) = .273, *p* = .005, *d* = .821).

The first hypothesis was fully supported by these findings. The second hypothesis predicted that there would be a positive relationship between perceived availability of on-campus dining services and disordered eating attitudes and behaviors in university students.

A Pearson correlation coefficient was computed to assess the linear relationship between perceived availability of nutritional services and disordered eating behaviors. There was a positive correlation between the variables (*r*(86) = .289, *p* = .003, *d* = .774).

A Pearson correlation coefficient was also computed to assess the relationship between availability of nutritional services and disordered eating attitudes. Again, a positive relationship was identified (*r*(86) = .235, *p* = .014, *d* = .823).

Therefore, the second hypothesis was fully supported.

There was no statistically significant correlation between first-year dependence on on-campus dining services and risk behaviors related to eating disorders or disordered eating attitudes in university students.

Based on these findings, the quality of nutrition provided and the availability of campus dining services impacted university students’ eating attitudes and behaviors rather than inherent dependence itself.

## 5. General Discussion

The netnography conducted laid the foundation and provided justification for the second portion of this research project because it allowed the researcher to observe the subjective experiences of university students regarding campus dining services from a relatively distant perspective while identifying convergent themes represented in the categories above.

Both H1 and H2 were supported by findings indicating statistically significant positive relationships between perceived availability of services and disordered eating attitudes and behaviors, as well as overall perceived quality (satisfaction) with nutritional services and disordered eating attitudes and behaviors.

These findings indicate that the availability and accessibility of university dining services have statistically significant positive relationships with disordered eating attitudes and behaviors in university students. This means that as university students reported increasingly negative perceptions regarding the availability of dining services, they also demonstrated increasingly negative eating behaviors and disordered eating attitudes.

These findings answer research questions two and three regarding the significance of the relationship between campus dining services and disordered eating attitudes and behaviors in university students.

With regard to RQ1, satisfaction was positively correlated with perceived quality of nutrition provided, and the availability of campus dining services impacted university students’ eating attitudes and behaviors rather than inherent dependence itself.

These findings appear consistent with **?**’s argument that increasing incidences of eating disorders may be attributed to changes in the food environment. This interpretation is also consistent with Huber et al. (2021)’s findings that disruptions caused by the COVID-19 pandemic significantly impacted food procurement and dietary habits among young adults.

Significant disruptions to an individual’s food environment may therefore produce substantial consequences for already vulnerable populations. Future research should investigate mitigating factors associated with major disruptions and transitions in the food environment. Additionally, future work should continue focusing on university students due to their increased susceptibility to disordered eating behaviors (DEBs) and disordered eating attitudes (DEAs).

The relationship between perceived availability of nutritional services and disordered eating pathology also aligns with Becker et al. (2019)’s findings that food insecurity may mimic the psychological effects associated with eating disorders.

Student health should remain a priority for university officials, and nutritional education programs represent an important step toward improving student well-being.

Health and nutrition education is especially important for university students because it may help modify, mitigate, and prevent maladaptive eating habits and behaviors while reducing the risk of illness, injury, and mortality, according to the US Department of Health and Human Services (2010). Such programs promote healthier eating attitudes, nutritional knowledge, and eating behaviors among university students (Eun-Jeong and Natalie, 2009).

Students should therefore be encouraged to participate in educational opportunities promoting healthy lifestyles and proper nutritional awareness.

According to Heinberg (2021), the starved brain associated with eating disorders, particularly anorexia nervosa, perpetuates irrational and maladaptive cognitive processes. This mechanism functions as a positive feedback loop that intensifies as the disorder progresses.

Understanding cognitive processes associated with perceptions of food availability and nutritional quality is therefore crucial for alleviating disordered eating attitudes and behaviors within vulnerable populations.

The food environment plays a substantial role in eating pathology, and understanding how environmental factors influence eating behaviors is essential for developing a more comprehensive understanding of disordered eating etiology.

This phenomenon, combined with Christensen et al. (2021)’s findings that 47.6% of individuals experiencing food insecurity screened positive for an eating disorder, highlights the importance of understanding extraneous environmental variables influencing the development of disordered eating behaviors and attitudes in university students.

Given the substantial number of university students in the United States who experience food insecurity, these findings demonstrate the severity and significance of this issue.

## 6. Limitations and Future Directions

There are several limitations to this study that should be acknowledged. First, the sample size was relatively small, which may have affected the results. Future research should therefore aim to increase sample size and statistical power. Second, the data were self-reported, suggesting that future investigations should attempt to incorporate observational and behavioral measures. Finally, this correlational study utilized convenience sampling, with a significant portion of the sample being female (85%). Future research should therefore attempt to replicate these findings using samples that are more representative of broader student demographics and sex distributions.

To reduce the potential influence of confirmation bias in interpreting the findings, the second author assisted in the analysis and interpretation of the results.

This study provides a foundation for future research examining the relationship between perceived quality of food, availability of nutritional services, and university students’ eating attitudes and behaviors. Additionally, longitudinal studies would be beneficial in establishing causal relationships.

Because a portion of the participants were recruited through the university’s honors program, future research may benefit from specifically investigating high-achieving individuals and their eating pathology.

Qualitative methodologies may also be useful for exploring students’ lived experiences with disordered eating attitudes and behaviors in greater depth.

There are several additional avenues for future research in this area. One possible direction would involve conducting larger-scale studies with more diverse populations in order to further examine the impact of perceived nutritional service quality and availability across different environments and demographic groups.

Additionally, it may be beneficial to investigate the long-term impact of these phenomena to determine whether the observed effects persist over time.

Another important direction for future research would involve exploring the underlying mechanisms explaining why these relationships occur, including environmental changes and communication surrounding nutritional services through technological and social media platforms.

This could involve more in-depth analyses of specific components associated with campus dining services while examining potential mediating variables involved in the etiology of disordered eating behaviors, particularly communication-centered promotion of nutritional services upon which individuals rely.

The findings of this study also possess several practical implications for practitioners in educational, organizational, and community settings. Specifically, the findings may be useful in schools, workplaces, and community organizations to help improve eating attitudes and behaviors by changing how nutritional services are communicated about, thereby reinforcing more positive perceptions of food services.

The findings additionally possess several practical implications for campus dining services and university health systems. First, the study highlights the importance of providing highquality and diverse dining options to meet students’ nutritional needs. Campus dining services should also prioritize the accessibility and availability of food services, particularly for students with limited access to off-campus food options.

Providing education and resources supporting healthy eating habits may additionally help prevent disordered eating attitudes and behaviors among university students.

Finally, this study emphasizes the importance of continued research and evaluation of campus dining services and their impact on student health and well-being.

Specifically, nutritional education programs designed to increase awareness of healthy eating habits while reinforcing healthy eating intentions through mindfulness techniques may prove especially beneficial (Krishnan and Zhou, 2019). Krishnan and Zhou (2019) found that social media engagement regarding healthy eating attitudes and behaviors facilitated the maintenance of healthy nutritional behaviors. Thus, increased social media engagement in this context may reinforce healthy eating practices.

The incorporation of multiple dimensions of social support and social activity contributes to a centralized social influence construct (Krishnan and Zhou, 2019) that may have substantial potential for integration into broader systemic interventions targeting disordered eating behaviors (DEBs) and disordered eating attitudes (DEAs).

The food provided by universities is frequently characterized by high levels of sugars, fats, and carbohydrates while remaining relatively low in nutritional value. University students therefore need greater awareness regarding the healthiest and most advantageous food options available to them (Whybird et al., 2022).

Many universities do not provide nutrition labels on cafeteria-style food offerings. Ingredients and dietary information should therefore be listed alongside food items to help students make healthier nutritional decisions (Francis and Eleanor, 2005).

The lack of nutritional labeling and dietary guidance represents another important avenue for future research, particularly regarding student perceptions of nutritional service quality and the relationship between those perceptions and disordered eating attitudes and behaviors.

Additionally, implementing nutritional awareness programs within university cafeterias may produce substantial long-term benefits for students’ health behaviors and potentially reinforce lifelong healthy eating practices.

## Data Availability Statement

The data supporting this study’s findings are openly available through Science Data Bank at: https://doi.org/10.57760/sciencedb.13426

## Geolocation Information

This research study was conducted in Vermillion, South Dakota, United States. Vermillion is a city located in Clay County, South Dakota, on the east bank of the Missouri River approximately 45 miles south of Sioux Falls, South Dakota. Vermillion is home to the University of South Dakota, the state’s flagship public university.

## Appendix

All Yik Yak posts from August 2021 to December 2022 were utilized in the thematic coding of qualitative data to comprise Themes 1 and 2.

### 6.1. First Semester Provider: Aramark

“Finals coming up [emoji] my ED making a reappearance = a recipe for disaster.”

“Can my ED go away this is just annoying now.”

“Comment/upvote if you believe that campus dining led you to develop an eating disorder or worsened your already existing ED.”

“I eat a spoonful of Nutella a day.” “So, it wasn’t just me.”

“I never eat… It’s so bad.”

“Just found a piece of a glove in my [Mexican fastfood restaurant] burrito bowl.”

#### 6.2. Second Semester Provider: Sodexo

“Is the [campus dining service] food giving anyone else diarrhea?”

“Absolutely.”

“Hershey squirts are tough [right now].”

“Anyone else have absolutely terrible shits from [campus dining service] food.”

“Straight from the bowels of hell…” “Anything I eat in there I swear to god.”

“The [campus dining service] food gives me the shits 0/10.”

“Swear to god [Mexican fast-food restaurant] gave me food poisoning.”

“Anyone else get sick EVERY time they eat [Mexican restaurant] food?”

“Hair in my [Mexican fast-food restaurant].”

“The [student dining service] is OBLITERATING my stomach.”

“The food in the YUC makes me puke.”

“Upvote if the [campus dining services Mexican restaurant] has given you food poisoning.”

“[Campus dining service] food got anyone else shitting 2–3 times a day???????”

“My stomach can’t handle [campus dining service] food anymore… I’m in shambles.”

“[Mexican fast-food restaurant] got my ass in the bathroom every 5 minutes I swear.”

“How the fuck is the cafeteria already closed at 6:27pm???”

“I went at 5:30 and it was closed.”

“Finals coming up [emoji] my ED making a reappearance = a recipe for disaster”

“Can my ED go away this is just annoying now”

“Comment/upvote if you believe that campus dining led you to develop an eating disorder or worsened your already existing ED.”

“I eat a spoonful of Nutella a day.” “So, it wasn’t just me”

“I never eat… It’s so bad.”

